# Monkeypox Post COVID19: Knowledge, Worrying, and Vaccine Adoption of the Arabic General Population

**DOI:** 10.1101/2022.12.20.22283750

**Authors:** Sarya Swed, Haidara Bohsas, Hidar Alibrahim, Amine Rakab, Wael Hafez, Bisher Sawaf, Mohammed Amir Rais, Ahmed Sallam Motawei, Ahmed Aljabali, Shiekh shoib, Ismail Atef Ismail Ahmed Ibrahim, Sondos Hussein Ahmad Almashaqbeh, Ebrahim Ahmed Qaid Shaddad, Maryam AlShaqsi, Ahmed abdelrahman, Sherihan fathey, René Hurlemann, Mohamed Elsayed, Bijaya Kumar Padhi, Ranjit Sah

## Abstract

**Background:** The outbreak of monkeypox was designated a global public health emergency by the World Health Organization on July 23, 2022. There have been more reported 60000 cases worldwide, most of which are in places where monkeypox has never been seen due to the travel of people who have the virus. This research aims to evaluate the Arabic general population on monkeypox disease, fears, and vaccine adoption after the WHO proclaimed a monkeypox epidemic and to compare these attitudes to those of the COVID-19 pandemic.

**Methods:** This cross-sectional study was performed in some Arabic countries (Syria, Egypt, Qatar, Yemen, Jordan, Sudan, Algeria, and Iraq) between August 18 and September 7, 2022 to examine the Arabic people perspectives on monkeypox disease, fears, and vaccine adoption and to compare these attitudes to those of the COVID-19 pandemic. The inclusion criteria were the general public residing in Arabic nations and older than 18. This questionnaire has 32 questions separated into three sections: sociodemographic variables, prior COVID-19 exposure, and COVID-19 vaccination history. The second portion assesses knowledge and anxieties about monkeypox, while the third section includes the generalized anxiety disorder (GAD7) scale. Logistic regression analysis were performed to compute the adjusted odds ratios (aOR), and their confidence intervals (95%CI) using STATA (version 17.0)

**Results:** A total of 3665 respondents from 17 Arabic countries were involved in this study. Almost two third (n= 2427, 66.2%) of participants expressed more worried about COVID -19 than monkeypox diseases. Regarding the major cause for concern about monkeypox, 39.5% of participants attributed their anxiety they or a member of their family may contract the illness, while 38.4% were concerned about another worldwide pandemic of monkeypox. According to the GAD 7 score, 71.7% of respondents showed very low anxiety toward monkeypox. 43.8% of the participants scored poor levels of knowledge about monkeypox disease. Participants with previous COVID-19 infection showed greater acceptance to receive the monkeypox vaccine 1.206 times than those with no previous infection. A higher concern for the monkeypox than COVID-19 was shown by the participants who perceived monkeypox as dangerous and virulent 3.097 times than those who didn’t. Participants who have a chronic disease (aOR: 1.32; 95%CI: 1.09-1.60); participants worried about monkeypox (aOR: 1.21; 95%CI: 1.04-1.40); and perceived monkeypox as a dangerous and virulent disease (aOR: 2.25; 95%CI: 1.92-2.65); and excellent knowledge level (aOR: 2.28; 95%CI: 1.79-2.90) have emerged as significant predictors.

**Conclusion:** Our study reported that three fourth of the participants were more concerned about COVID-19 than monkeypox disease. As well, most of the participants have inadequate levels of knowledge regarding monkeypox disease. Hence immediate action should be taken to address this problem. Consequently, it is crucial to learn about monkeypox and spread information about its prevention.

## Introduction

The outbreak of monkeypox was declared a global public health emergency condition by the World Health Organization on July 23, 2022. The majority of the reported cases were from Europe [1]. Since the beginning of the COVID-19 epidemic, nearly 2 million fatalities have occurred worldwide. Extensive research has shown a correlation between COVID-19 and mental health problems such as anxiety, depression, and mental distress. This association increased as the worldwide number of COVID-19 cases and fatalities increased [2]. The monkeypox virus is a double-stranded DNA virus that can spread from animals to humans. It is in the genus Orthopoxvirus and the family Poxviridae [3]. There are about 21504 cases in the US, but only 35 cases have been reported from Arabic countries [4]. Multiple animal hosts for this virus have been discovered, including rope squirrels, tree squirrels, Gambian pouched rats, dormice, and many kinds of monkeys [5]. The predicted incubation time for monkeypox ranges from 5 to 21 days. Initial illness signs are vague, including fever, chills, headaches, lymphadenopathy, back pain, myalgia, and eventually, the rash appears [6]. This disease has been linked to several complications, including bacterial superinfection of the skin, pneumonia, encephalitis, sepsis, and death [7]. Psychiatric and neurological manifestations might also occur [8]. In most situations, monkeypox infections heal without medical treatment. Oral or intravenous rehydration is advised to maintain hydration levels in patients with digestive symptoms such as vomiting and diarrhea. Several antiviral drugs, including tecovirimat, brincidofovir, and cidofovir, have shown effective against the monkeypox virus [9]. There has been evidence of the vaccine’s effectiveness in protecting against the monkeypox virus. Modified vaccinia Ankara and ACAM2000 are the two vaccines that have been established [10]. Recent research have shown that the general public is more concerned about COVID-19 than about monkeypox. Deaths from monkeypox have not been documented in areas where the disease is not naturally present, lending credence to the idea that the current outbreak’s clinical repercussions are less than those seen in endemic areas [11, 12]. However, there is still a gap in the literature of assessment of the knowledge and the psychological behaviors toward monkeypox illness and whether or if there is a more significant worry about it as the number of recorded cases increases. This research aims to evaluate the Arabic people perspectives on monkeypox disease, fears, and vaccine adoption after the WHO proclaimed a monkeypox epidemic and to compare these attitudes to those of the COVID-19 pandemic.

## Methods

### Study Design

A cross-sectional online study was performed in 17 Arabic countries (Syria, Egypt, Qatar, Yemen, Jordan, Sudan, Algeria, Iraq, and other countries) between August 18 and September 7, 2022 to examine the Arabic people perspectives on monkeypox disease, fears, and vaccine adoption and to compare these attitudes to those of the COVID-19 pandemic. Participants were required to be individuals of the general public residing in Arabic nations and older than 18 years. Medical professionals, students, and staff were not eligible for participation. All participants were informed of the study’s goals, the researchers’ identities, their ability to opt-out of participation, the confidentiality and security of their data, and the importance of providing all requested information. This questionnaire was constructed using a complete, verified scale based on previous research [11, 13]. The survey was then translated from English into Arabic for the respondent’s comprehension by a native translator. Concerned about security, a Google form survey was developed and distributed over social media sites such as Facebook, WhatsApp, and Telegram. As well, in the individual governorates, retail malls, parks, public squares, and other public meeting areas were accessible for data collection, as were face-to-face interviews.

#### Sample size calculation

Depending on the Arab population count (https://data.worldbank.org/indicator/SP.POP.TOTL?locations=1A), and using Calculator.net (https://www.calculator.net/sample-size_calculator.html), we conducted a statistical power analysis for sample size calculation using a population proportion of 50%, a margin of error of 0.05, and a degree of confidence of 99%, so the minimum sample size was 385. The overall sample size was 3822 participants.

#### Measures

This questionnaire has 32 questions separated into three sections: sociodemographic factors, prior COVID-19 exposure, and COVID-19 vaccination history. The second portion assesses knowledge and anxieties about monkeypox, while the third section includes the Generalized Anxiety Disorder (GAD7) scale towards monkeypox virus [14].

#### Sociodemographic characteristics

This section contains six questions regarding the participant’s sociodemographic characteristics, including age, gender, place of residence, marital status, economic status, and chronic illness status (hypertensive, diabetes.etc.). This section also includes three questions about participants’ prior exposure to COVID-19, including whether or not they have ever been infected with the virus, whether or not they took preventative measures against contracting the disease during the COVID-19 pandemic, and whether or not they are more concerned about a possible outbreak of monkeypox than COVID-19 epidemic.

#### knowledge and concerns about monkeypox

There are 11 multiple-choice questions here on the monkeypox virus that participants may answer yes, no, or not sure to gauge their level of familiarity with the Monkeypox. Participants were asked whether they believed that monkeypox was caused by a virus or bacteria, whether it could be spread from person to person, whether its symptoms were similar to those of smallpox, whether its rash and papules were indicative of the disease, and whether antibiotics were effective against the virus. Extensive prior research uses of this instrument attest to its reliability [13]. There are also three questions in this section that assess how worried people are about monkeypox and related issues (such as whether or not the virus is serious enough to warrant taking precautions to avoid spreading it and whether or not people should be quarantined worldwide). In addition, this section includes two questions to assess participant willingness to get the monkeypox vaccination and to identify the most probable receivers. This question was extracted from a previously verified survey [11].

#### Generalized Anxiety Disorder (GAD) toward monkeypox

This tool consists of seven validated questions to assess the GAD of participants towards the monkeypox virus [14]. In this tool, respondents were asked to assess the frequency with which they had experienced symptoms such as worry, concern, restlessness, impatience, and dread during the previous two weeks. We gave values between 0 and 3 to the following frequency categories: never, sometimes, often, and very frequently. GAD7 scores were tallied and classified as minimal (0–4), mild (5–9), moderate (10–14), and severe (0–14). (15–21).

##### Pilot study

We administered the questionnaire to 50 Arabic individuals selected randomly to ensure its validity and readability, which we detected high levels of internal consistency (Cronbach’s alpha varied from 0.712 to 0.861).

Ethical Consideration: Ethical Approval is taken from Syrian Arab Republic, Aleppo city, Faculty of Medicine, Aleppo University. The IRB approval number is SA-AL 4853B

The Syrian Ethical Society for Scientific Research provided its stamp of approval (SA-AL4853B). Furthermore, at least one ethical approval was taken from each inquired country in our study. Participants were given a Google Survey URL and asked to confirm their agreement to participate in the study on the first page of the survey. There is a wealth of background reading for participants to peruse before they fill out the questionnaire on the next page and complete this survey within 5-12 minutes. The responses were collected and kept in an encrypted internet database.

#### Statistical analysis

The statistical data analysis was done using the IBM SPSS statistical analysis tool (28). Means and standard deviations were used in the process of describing continuous variables, whilst frequencies and percentages were utilized in the process of describing categorically measured variables. The Multivariate Binary Logistic Regression Analysis was used so that investigators could determine what variables could be responsible for people’s concerns over monkeypox and their willingness to be vaccinated against the disease. An odds ratio (OR) with accompanying 95% confidence intervals was used to represent the connection between predictors and the outcome dependent variables in the multivariate Logistic Binary regression analysis. The statistical Alpha significance value of 0.050 was being used.

## Results

The demographic variables of the respondents are summarized in Table 1. A total of 3772 responses from 17 Arabic countries were collected in this study, however the final eligible sample size for data analysis after omitting the missing data was 3665. The highest proportion of the inquired participants was from Yemen (n = 689, 18.8 %), followed by Syria (n=394,10.8%). More than half of the participants were female (2187, 59.7 %). Most respondents (n=3016, 82.3 %) were 18 – 34 years old. Nearly two third of study participants were single (2249, 61.4%). Regarding the economic status, 56.1 % (n= 2055) of respondents had moderate economic condition, while 30.6 % (n = 1123) had good economic condition. The majority of the participants (84.0 %, n = 3078) had no history of chronic disease.

**Table 1.**
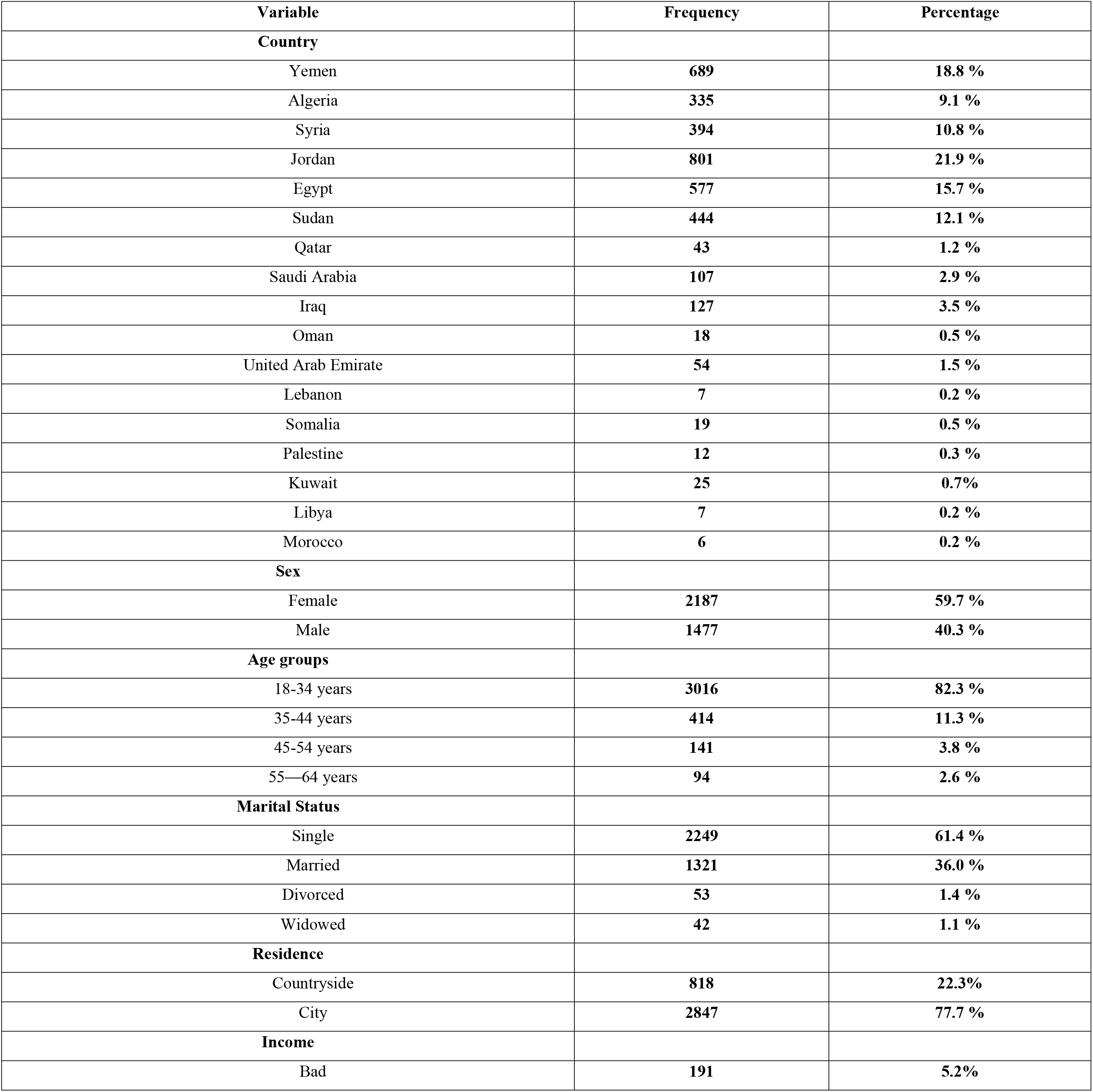
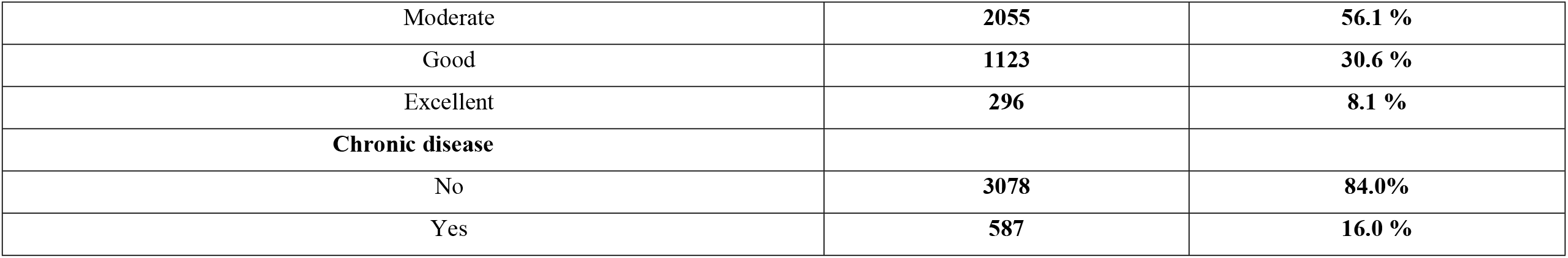
Sociodemographic characteristics of participants (**N=3665**)

Regarding table 2, 45.8% (n= 1679) of participants reported a previous infection by COVID -19 disease; also, about half of the participants (51.9%, n= 1903) reported a Medium commitment during the COVID-19 pandemic. Almost two third (n= 2427, 66.2%) of participants expressed more worried about COVID -19 than monkeypox diseases. Regarding the major cause for concern about monkeypox, 39.5% of participants attributed their anxiety they or a member of their family may contract the illness, while 38.4% were concerned about another worldwide pandemic of monkeypox. 69.9 % of respondents agreed that monkeypox is a dangerous and virulent illness that requires respiratory and contact precautions. According to the GAD 7 score, 71.7% of respondents showed very low anxiety toward monkeypox. However, (n= 1604, 43.8%) scored poor levels of knowledge regarding monkeypox disease (Table 2).

**Table 2.** Respondents’ attitudes, perceptions, and beliefs about Monkeypox disease (N=3665).

| Variable | Frequency | Percentage |
| --- | --- | --- |
| Have you been affected by the COVID-19 disease? |  |  |
| No | 1986 | 54.2 % |
| Yes | 1679 | 45.8 % |
| Participant's and family members compliance with COVID-19 pandemic precautions |  |  |
| Rarely committed | 602 | 16.4 % |
| Medium commitment | 1903 | 51.9 % |
| Always committed | 1160 | 31.7 % |
| Worry from Monkeypox compared to COVID-19 |  |  |
| Much worried with the COVID-19 | 2427 | 66.2 % |
| More worried with Monkeypox | 1238 | 33.8 % |
| Main reasons for your worry from Monkeypox disease |  |  |
| Self and family worry of Monkeypox infection | 1447 | 39.5 % |
| Worry from another worldwide pandemic | 1409 | 38.4 % |
| Worried that Monkeypox might surge to cause a national lockdown | 400 | 10.9 % |
| Concerned that an international flight suspension happens | 79 | 2.2 % |
| Other worries | 330 | 9.0 % |
| Do you perceive Monkeypox as a dangerous and virulent disease that calls for respiratory and contact precautions? |  |  |
| No | 1102 | 30.1 % |
| Yes | 2563 | 69.9% |
| GAD7 score, mean (SD) | 3.15 (4.46) | - |
| GAD7 score levels |  |  |
| Very low Anxiety | 2627 | 71.7 % |
| Mild Anxiety | 621 | 16.9 % |
| Moderate Anxiety | 321 | 8.8 % |
| High Anxiety | 96 | 2.6 % |
| Overall Monkeypox Disease Knowledge score, mean (SD). Maximum score = 11 | 3.99 (2.87) | - |
| Overall Monkeypox Disease Knowledge score levels |  |  |
| Poor | 1604 | 43.8 % |
| Good | 1668 | 45.5 % |
| Excellent | 393 | 10.7 % |

**Table 3.** Multivariate Binary Logistic Regression Analysis of people’s odds of agreement to vaccinate against Monkeypox Disease, (Agree versus Disagree).

| Predictor | Adjusted Odds ratio | 95% Confidence Interval |  | P value |
| --- | --- | --- | --- | --- |
|  |  | Lower | Upper |  |
| Sex |  |  |  |  |
| Female – Male | 0.857 | 0.731 | 1.005 | 0.058 |
| Age Groups |  |  |  |  |
| 35-44 – 18-34 years | 0.715 | 0.555 | 0.922 | 0.01 |
| 45-54 – 18-34 years | 0.612 | 0.408 | 0.918 | 0.018 |
| 55—64 – 18-34 years | 0.843 | 0.527 | 1.348 | 0.475 |
| Marital Status |  |  |  |  |
| Married – Single | 0.855 | 0.718 | 1.019 | 0.08 |
| Divorced – Single | 0.6 | 0.331 | 1.091 | 0.094 |
| Widowed – Single | 0.952 | 0.481 | 1.885 | 0.887 |
| **Residence** |  |  |  |  |
| City – Countryside | 0.81 | 0.678 | 0.968 | 0.021 |
| **Income** |  |  |  |  |
| Moderate – Bad | 1.159 | 0.834 | 1.611 | 0.38 |
| Good – Bad | 1.278 | 0.905 | 1.805 | 0.163 |
| Excellent – Bad | 2.069 | 1.371 | 3.122 | < .001 |
| **Chronic Disease** |  |  |  |  |
| Yes – No | 1.193 | 0.976 | 1.458 | 0.085 |
| **Previous COVID-19 infection** |  |  |  |  |
| Yes – No | 1.206 | 1.031 | 1.411 | 0.019 |
| **Participant and family members compliance** |  |  |  |  |
| Medium commitment – Rarely committed | 0.871 | 0.711 | 1.066 | 0.181 |
| Always committed – Rarely committed | 1.012 | 0.804 | 1.275 | 0.916 |
| **Worry from Monkeypox compared to COVID19** |  |  |  |  |
| More worried with Monkeypox – Much worried with the COVID-19 | 1.111 | 0.948 | 1.302 | 0.193 |
| **Main reasons for your worry from Monkeypox disease** |  |  |  |  |
| Worry from another worldwide pandemic – Self and family worry of Monkeypox infection | 0.949 | 0.809 | 1.115 | 0.526 |
| Worried that Monkeypox might surge to cause a national lockdown – Self and family worry of Monkeypox infection | 0.74 | 0.579 | 0.945 | 0.016 |
| Concerned that an international flight suspension happens – Self and family worry of Monkeypox infection | 0.56 | 0.335 | 0.935 | 0.027 |
| Other worries – Self and family worry of Monkeypox infection | 0.461 | 0.349 | 0.607 | < .001 |
| **Do you perceive Monkeypox as a dangerous and virulent** |  |  |  |  |
| Yes – No | 1.889 | 1.591 | 2.243 | < .001 |
| **Total Knowledge Score** | 1.099 | 1.07 | 1.128 | < .001 |
| **Total GAD Score** | 1.045 | 1.028 | 1.064 | < .001 |

Ten out of fourteen predicted variables were statistically significant associated with more willingness to vaccinate against monkeypox (p < 0.05). Respondents aged between 35-44 years had fewer odds of willingness to receive the monkeypox vaccine compared to the 18-34 age group (OR= 0.715). In comparison to those who were single, married respondents were projected to have less acceptance of the monkeypox vaccination (OR= 0.855). Participants with excellent income were more likely 2.069 times to receive the vaccine than those with bad income. Participants with previous COVID-19 infection showed greater acceptance to receive the monkeypox vaccine 1.206 times than those with no previous infection. Regarding the main reasons for worry about Monkeypox disease, participants who expressed worry that Monkeypox might surge to cause a national lockdown, who were concerned that an international flight suspension would happen, and who reported other reasons of worry had a significant drop in vaccination acceptability 0.016, 0.027, < .001 times respectively than those expressed concerned they or a member of their family could contract the illness. A higher agreement for the monkeypox vaccine was expected by the participant who perceived monkeypox as dangerous and virulent 1.889 times more than those who didn’t.

Eight out of fourteen predictor factors were significantly linked with greater worry from Monkeypox than COVID-19 (P-value< 0.05). Females were more likely to be concerned about Monkeypox 1.379 times more than males. Participants aged 35-44 had low odds of worry about Monkeypox compared to 18-34. Low worries were predicted among participants with chronic disease 0.731 times more than those without. Regarding the main reasons for worry about Monkeypox disease, participants who expressed worry that Monkeypox might surge to cause another worldwide pandemic had a significant drop in worry about Monkeypox 0.723 times than those expressed concern they or a member of their family could contract the illness. A higher concern for the Monkeypox than COVID-19 was shown by the participant who perceived Monkeypox as a dangerous and virulent 3.097 times than those who didn’t.

Table 5 depicts the findings of multiple logistic regression analysis for the association between intention to vaccinate and socio-behavioral characteristics of study participants. Participants who have a chronic disease (aOR: 1.32; 95%CI: 1.09-1.60); participants worried about monkeypox (aOR: 1.21; 95%CI: 1.04-1.40); and perceived monkeypox as a dangerous and virulent disease (aOR: 2.25; 95%CI: 1.92-2.65); and excellent knowledge level (aOR: 2.28; 95%CI: 1.79-2.90) were emerged as significant predictors.

**Table 4.**
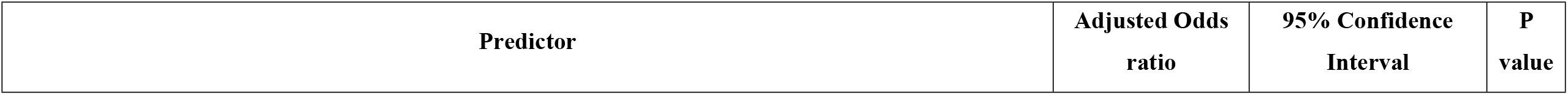
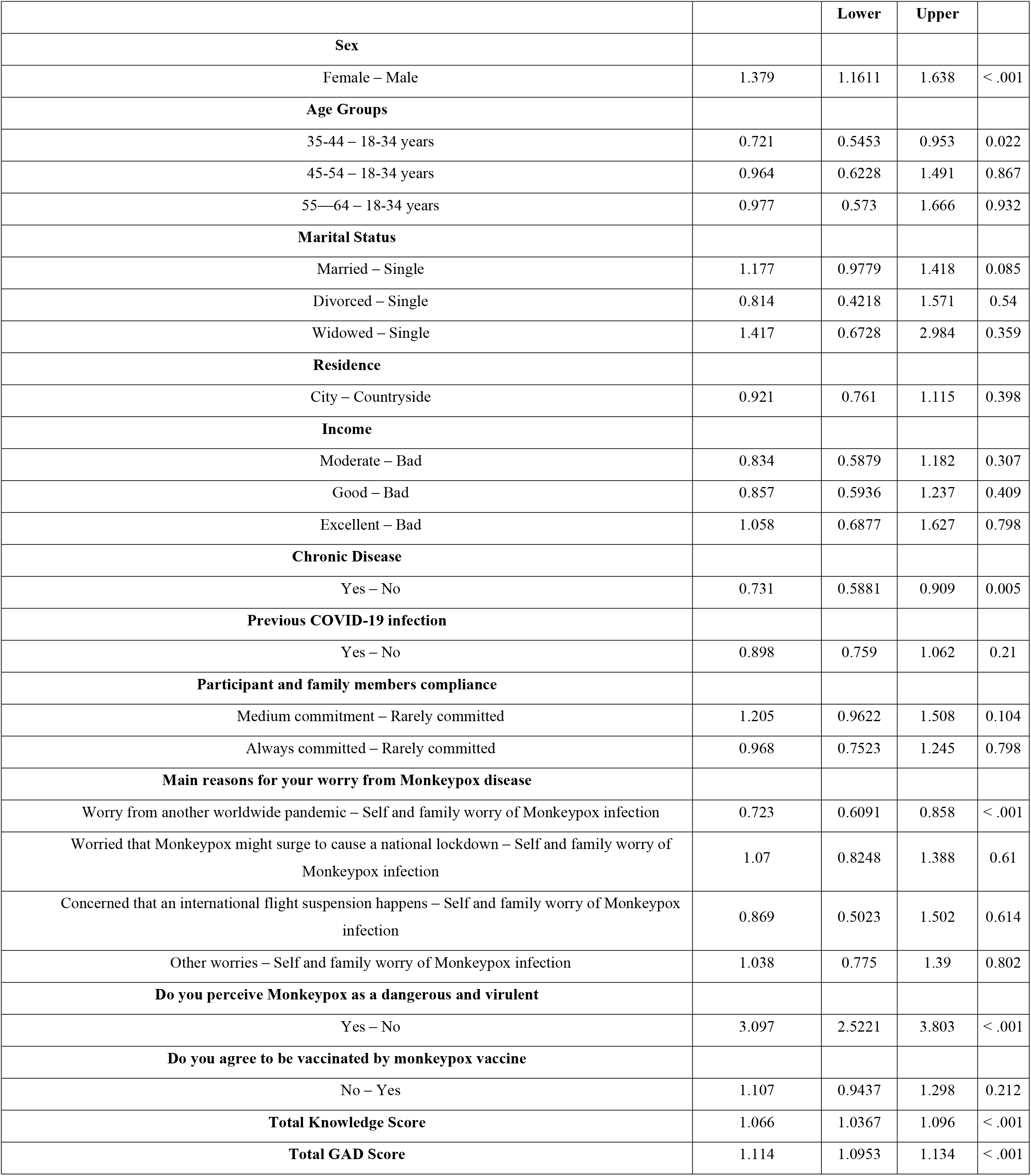
Multivariate Binary Logistic Regression Analysis of respondents’ odds of higher worry level from Monkeypox disease compared to COVID-19.

| Predictor | Adjusted Odds ratio | 95% Confidence Interval | P value |
| --- | --- | --- | --- |

|  |  |  |  |  |
| --- | --- | --- | --- | --- |
|  |  | Lower | Upper |  |
| Sex |  |  |  |  |
| Female – Male | 1.379 | 1.1611 | 1.638 | < .001 |
| Age Groups |  |  |  |  |
| 35-44 – 18-34 years | 0.721 | 0.5453 | 0.953 | 0.022 |
| 45-54 – 18-34 years | 0.964 | 0.6228 | 1.491 | 0.867 |
| 55—64 – 18-34 years | 0.977 | 0.573 | 1.666 | 0.932 |
| Marital Status |  |  |  |  |
| Married – Single | 1.177 | 0.9779 | 1.418 | 0.085 |
| Divorced – Single | 0.814 | 0.4218 | 1.571 | 0.54 |
| Widowed – Single | 1.417 | 0.6728 | 2.984 | 0.359 |
| Residence |  |  |  |  |
| City – Countryside | 0.921 | 0.761 | 1.115 | 0.398 |
| Income |  |  |  |  |
| Moderate – Bad | 0.834 | 0.5879 | 1.182 | 0.307 |
| Good – Bad | 0.857 | 0.5936 | 1.237 | 0.409 |
| Excellent – Bad | 1.058 | 0.6877 | 1.627 | 0.798 |
| Chronic Disease |  |  |  |  |
| Yes – No | 0.731 | 0.5881 | 0.909 | 0.005 |
| Previous COVID-19 infection |  |  |  |  |
| Yes – No | 0.898 | 0.759 | 1.062 | 0.21 |
| Participant and family members compliance |  |  |  |  |
| Medium commitment – Rarely committed | 1.205 | 0.9622 | 1.508 | 0.104 |
| Always committed – Rarely committed | 0.968 | 0.7523 | 1.245 | 0.798 |
| Main reasons for your worry from Monkeypox disease |  |  |  |  |
| Worry from another worldwide pandemic – Self and family worry of Monkeypox infection | 0.723 | 0.6091 | 0.858 | < .001 |
| Worried that Monkeypox might surge to cause a national lockdown – Self and family worry of Monkeypox infection | 1.07 | 0.8248 | 1.388 | 0.61 |
| Concerned that an international flight suspension happens – Self and family worry of Monkeypox infection | 0.869 | 0.5023 | 1.502 | 0.614 |
| Other worries – Self and family worry of Monkeypox infection | 1.038 | 0.775 | 1.39 | 0.802 |
| Do you perceive Monkeypox as a dangerous and virulent |  |  |  |  |
| Yes – No | 3.097 | 2.5221 | 3.803 | < .001 |
| Do you agree to be vaccinated by monkeypox vaccine |  |  |  |  |
| No – Yes | 1.107 | 0.9437 | 1.298 | 0.212 |
| Total Knowledge Score | 1.066 | 1.0367 | 1.096 | < .001 |
| Total GAD Score | 1.114 | 1.0953 | 1.134 | < .001 |

**Table 5:** Multiple logistic regression analysis for the association between intention to vaccinate and socio-behavioral characteristics of study participants (N= 3664)

| Variable | aOR [95% CI] | P-value |
| --- | --- | --- |
| **Age (Reference: 18-34)** |  |  |
| 35-44 | 0.65 [0.51, 0.83] | 0.00 |
| 45-54 | 0.59 [0.40, 0.87] | 0.01 |
| 55—64 | 0.86 [0.54, 1.36] | 0.51 |
| **Gender (Reference: male)** |  |  |
| Female | 1.08 [0.93, 1.24] | 0.31 |
| **Marital Status (Reference: Single)** |  |  |
| Married | 0.88 [0.74, 1.03] | 0.11 |
| Divorced | 0.76 [0.42, 1.36] | 0.35 |
| Widowed | 0.99 [0.51, 1.93] | 0.97 |
| **Residence (Reference: Countryside)** |  |  |
| City | 0.76 [0.65, 0.90] | 0.00 |
| **Chronic Disease (Reference: No)** |  |  |
| Yes | 1.32 [1.09, 1.60] | 0.00 |
| **Previous COVID19 Infection (Reference: No)** |  |  |
| Yes | 1.08 [0.94, 1.23] | 0.31 |
| **Participant's and family members compliance with COVID-19 pandemic precautions (Reference: Rarely committed)** |  |  |
| Medium commitment | 0.83 [0.69, 1.01] | 0.06 |
| Always committed | 0.88 [0.71, 1.08] | 0.22 |
| **Worry from Monkeypox compared to COVID-19 (Reference: Much worried with the COVID-19)** |  |  |
| More worried with Monkeypox | 1.21 [1.04, 1.40] | 0.01 |
| **Do you perceive Monkeypox as a dangerous and virulent disease that calls for res (Reference: No)** |  |  |
| Yes | 2.25 [1.92, 2.65] | 0.00 |
| **Knowledge of Monkeypox (Reference: Poor)** |  |  |
| Good | 1.62 [1.39, 1.87] | 0.00 |
| Excellent | 2.28 [1.79, 2.90] | 0.00 |

## Discussion

The recent spread of the global health pandemics is one of the major problems facing humanity and affecting the world in various scales of life [15]. COVID-19 pandemic during the past three years is the best example of the extent to which the humanity might be affected by such outbreaks. Accordingly, the recent announcement by the WHO of the possibility of facing the threat of a new global pandemic (i.e. Monkeypox (MPX) as a Public Health Emergency of International Concern) [16], in the midst of our confrontation with the COVID-19 pandemic, pushes the world to be vigilant and take the necessary precautionary measures to avoid the recurrence of the tragedy of the Corona-virus pandemic and other previous intractable pandemics. However, this caution may result in episodes of anxiety and depression of varying degrees among the mass of people that may repeat what they have experienced during the harsh conditions of COVID-19 pandemic [17-20]. In our cross sectional online survey, we assessed the level of anxiety among a sample of Arab population after the prevalence of Monkeypox virus (MPXV) in some Arab countries. Less than half 45.8% of the participants confirmed former COVID-19 infection history while slightly more than half 51.9% reported a medium commitment and average rules follow during the Covid-19 pandemic. This constant compliance with coronavirus measure control is crucial to be taken into consideration for possible continued education and awareness messages concerning the Monkey pox disease. Despite the increase of MPX cases in Arab countries [4], very low anxiety concerning this rising health problem was recorded among most of the participants (71.7%) according to GAD7 score. This may be attributed to the low level of knowledge about the disease among people, as we found that 43.8% of the participants recorded a poor level of knowledge about the disease. The small number of cases of monkeypox and the rarity of men who engage in male to male sex in Arabic-speaking nations may have had an impact on our findings by lowering people’s awareness of and fear of the disease. Accordingly, it is necessary to conduct awareness campaigns about its methods of transmission, prevention and management, for the public through the media and in schools at different stages. In addition, with the world still facing COVID-19 pandemic at the present time and since 2020, we compared the levels of public worry of the MPX epidemic with that of COVID-19 pandemic. We found that most of the participants (66.2%) expressed more worry about COVID-19 than MPX. However, more concern and worry about MPX was paid among some groups of the participants than their equivalents (e.g. females (OR=1.379), Participants aged 18-34, Participants without chronic disease (OR=1.368), more details in the results). Similar findings were reported by the COVID-19 Mental Disorders Collaborators in their large systematic review about the prevalence of major depressive disorders and anxiety disorders induced by COVID-19 pandemic[17]. But, the reasons for anxiety about MPX differ from those for COVID-19. We found that most of the anxiety in our study stemmed from their fear that they or a member of their family might be infected with MPXV, or worry from another worldwide pandemic and its consequences. In contrast, fear of international flights suspension was reported as the less worrying factor among the study population and, the major worries during the COVID-19 were from the high mortality rates and morbidities associated with the disease[17, 21]. Although there is not a specific vaccine that protects against MPXV, many orhtopoxvirus vaccines were found to cross-protect against MPXV by the antigenic similarity [22, 23]. Smallpox vaccines have great efficacy in prevention of MPX infection with clinical effectiveness of about 85%[24]. According to CDC, two smallpox vaccines (ACAM2000 AND JYNNEOS) are approved to be used in protection against both smallpox and MPX in the US [25]. Monkeypox vaccination can be given as either pre-exposure or post-exposure immunization [22, 24, 25]. While pre-exposure vaccination can reduce the risk of catching the infection, post-exposure vaccination within 4-14 days can ameliorate the symptoms of the disease and help the rapid recovery [24]. Based on our analysis, a large percentage of the respondents to our survey showed acceptance to MPXV vaccination. Thus, an effective measure that can help control the outbreak in the Arab world is the implementation of vaccination for high risk groups and the ring vaccination (i.e. vaccination of direct contacts of confirmed infected people [24, 26]. In light of this, the previous experience of COVID-19 pandemic stresses the importance of implementation of strong vaccination strategies. Based on Watson et.al analysis about the global impact induced by COVID-19 vaccination, millions of deaths were avoided after the vaccination with an estimated global reduction of 63% of the total deaths within the first year of vaccination [27]. With a major global event such the 2022 FIFA World Cup in Qatar approaching, strict precautions must be taken to avoid the spread of MPX and thus facing the risk of a new global pandemic beside the current COVID-19 one [28]. This might include: (1) raising the awareness and knowledge about the disease through social, media and educational campaigns, (2) conducting screening tests for the travelers before entering the country, (3) emphasizing the ability of the healthcare system to manage the situation, (4) the rapid diagnosis and control of any new cases, and (5) taking the MPXV vaccination strategy into consideration [29]. Finally, Ennab et al. and Sah et al. presented some of the commandments and lessons learned from COVID-19 pandemic, which we can benefit from to reduce the risk of facing a new health crisis [30, 31]. The concern and worry of the Arab population in the MENA region being average, reasonable and not exaggerated should not lead to a neglect that will result in a new pandemic, and yet Monkeypox infection appears to cause less mortality than Covid-19 infection, the symptoms tend to be more severe and difficult, moreover, the world health authorities are already weakened by the coronavirus pandemic which imposes a strong preparation for a probable second pandemic in the 21^st^ century caused by Monkeypox [21].

### Strengths and limitations

Although cross-sectional studies are inexpensive and can be easily accomplished, and analytical cross-sectional surveys may be used to study the relationship between a potential risk factor and a health result, the credibility of the assumptions that can be made about the relationship between a risk factor and a health result from this kind of research is limited. Furthermore, those who live in really distant areas, don’t have access to the internet, or are elderly won’t be able to participate in the research by completing the online survey. In our study, an investigator was assigned to each of the countries we asked about in order to continuously review the data collection procedure and omit the random and multiple auto-responses. As well, we did not include healthcare professionals or medical students.

## Conclusion

Concerns about new cases of Monkeypox in Arabic countries were lower than those for COVID-19. The two most significant determinants, the rarity of males who partake in male to male sex in Arabic-speaking regions and the low number of monkeypox cases, may have affected our results by reducing people’s knowledge of and fear of the virus. In conclusion, the historical experience of the COVID-19 pandemic highlights how essential it is to adopt effective vaccination and preventive policies.

## Data Availability

data is available with the first and corresponding authors, it can be provided on reasonable request

## Data collection group

Jarjees Abduljabar Sulaiman (University of duhok Iraq)

Hashem Altabbaa (University: Alexandria University-Egypt)

Albaraa Daradkeh (University: Alexandria University, Egypt)

Hebah Khaled Rababa (The university of jordan, Jordan)

ZUHAIR ANTAKLY (University of Aleppo, Faculty of Medicine, Syria)

BARA’AH AYED SULIEMAN AL ODAT (University of Jordan)

Dua Hassan Hassan Mohammed Abu-Ali (Sana’a University Faculty of Medicine)

Amatalkhaleq Hussein Hamoud Azzam (Sana’a Universit, Faculty of Medicine and Health Science, Sana’a)

WESSAL HASSAN ALI ABDALBAGI (Alneelain university, Khartoum _ Sudan)

Somaia Hafiz Ahmed (University of Science and Technology - Faculty of Medicine, Sudan)

Rais Mohammed Amir (Faculty of Medicine of Algiers, University of Algiers 1, Algeria)

Ayat Abdu Khalid Al-Mekhlafi (University of Jordan Yemen)

Nora Ra’ed Abdullah Atiah (Sana’a university faculty of medicine and health sciences, Yemen)

Mohamed Basyouni Elsayed Helal (Menoufia university faculty of medicine, Egypt)

ALHAMZA ALDARWIS (University: Damascus university faculty of medicine Country: Syria)

Duaa Bakdounes (Faculty of Medicine, Syrian Private University, Damascus, Syria)

## Ethics Statement

We confirm that written formal consent was obtained for each participant (The first question of the online survey).

